# Community and Socioeconomic Factors Associated with COVID-19 in the United States: Zip code level cross sectional analysis

**DOI:** 10.1101/2020.04.19.20071944

**Authors:** Avirup Guha, Janice M. Bonsu, Amit Kumar Dey, Daniel Addison

## Abstract

**Background:** Multiple reports have pointed towards involvement of community and socioeconomic characteristics of people in the United States may be associated with COVID-19 cases and deaths.

**Methods:** In this study, zip-code level data from 5 major metropolitan areas, was utilized to study the effect of multiple demographic & socioeconomic factors – including race, age, income, chronic disease comorbidity, population density, number of people per household on number of positive cases and ensuing death. Adjusted linear regression analysis using 13 to 16 such variables was performed.

**Results:** Overall, 442 zip codes reporting 93,170 positive COVID-19 cases and 138 zip codes reporting mortality ranging from 0 to 25 were included in this study. A multivariable linear regression model noted that 1% increase in the proportion of residents above the age of 65 years, proportion of African American residents, proportion of females, persons per household and population density of the zip code increased the proportion of positive cases by 0.77%, 0.23%, 1.64%, 1.83% and 0.46% respectively (P<0.01) with only population density remaining significant in zip codes with greater than median number of cases. In zips with greater than median number of deaths, no community/socio-economic factor contributed significantly to death.

**Conclusion:** This study gives early signals of gender, and racial inequalities while providing overwhelming evidence of how population density may contribute to an increase in the number of positive cases of COVID-19.

## Introduction

On March 11, 2020 the World Health Organization (WHO) declared the severe acute respiratory syndrome coronavirus 2 (SARS-CoV-2) a global pandemic (1). The impact of the novel coronavirus 2019 (COVID-19) has disrupted global markets, healthcare systems, and societies. The Johns Hopkins Coronavirus Resource Center, which tracks cases in real-time, has reported over 2,394,000 confirmed global cases, with over 164,000 deaths – nearly 35,000 in the United States alone (2). Economists liken the impact of this crisis on global societies to that at the end of World War II and estimate that it will take the United States (U.S.) and Eurozone three years to recover from COVID-19 (3).

With an increasing number of confirmed U.S. cases, COVID-19 is reminiscent of other infectious disease epidemics before it, with inequities in the prevalence of viral infection a growing concern. Historically, the HIV epidemic nearly exclusively impacted those who faced economic adversity and had lower levels of educational attainment (4). The relationship between neighborhood and area effects on health has been widely established (5). During this COVID-19 pandemic, reports on racial disparities have emerged, with reports of infection rates more than 3-fold higher in predominantly African American counties in the U.S. than in predominantly White counties (6). This is concerning given the potentially devastating consequences of this infection and the limited therapeutic options available.

Historically, many of these populations have shared a higher burden of chronic health factors linked with discrepancies in diseases outcomes. Yet, whether these factors actually affect COVID-19 related outcomes is largely unknown. In our study, we hypothesized that using zip code level data in 5 major metropolitan areas, we would observe that multiple socioeconomic factors – including race, age, income, chronic disease comorbidity, population density, number of people per household – impact the risk of COVID-19 infections.

## Methods

### Data Source

Authors collected data zip code level data on 4/11/2020 for the City of New York including all boroughs, Chicago, Richmond county of Detroit, Kings County of Seattle, Miami-Dade County. All data sources are publicly available and the corresponding URLs are listed in **supplemental Table 1**.

### Primary Outcome

The outcomes of interest for zip code level data for the various locations was the number of positive cases and reported causes of specific mortality.

### Covariates

The zip code level data was obtained from the censusreporter.org website for 446 zip codes spanning the areas listed above. Specific metrics collected were proportion of Whites, African Americans, and Hispanics, population density reported as people per square mile, proportion of females, median age in years, proportion of those older than 65 years, median household income in dollars, proportion of those below poverty line, persons per household, proportion of married, median house value in dollars, proportion of those with bachelor’s degree, and proportion of English speaking. Information on chronic disease from a uniform source was not available. Short-term complications for diabetes and chronic obstructive pulmonary disease (COPD) or asthma in older adults reported as an observed rate per 100,000 was collected for the New York zip codes. Diabetes-related hospitalizations and asthma related emergency room visits reported as per 10,000 population were collected for the Chicago zip codes. Temperature on March 15, 2020 was obtained from the National Oceanic and Atmospheric Administration (NOAA) website however this was city/county specific and only used for adjustment as warm (1) for Miami and cold (0) for other zip codes. Additionally, county level data for hypertension hospitalization per 1,000 population and cancer per 100,000 population was obtained from CDC website. This was dichotomized for those above and below the median of each metric and used only for adjustment. All the covariate sources are listed in **supplemental table 2**.

### Statistics

Zip code level data used total number of positive cases and deaths from COVID-19 as continuous variables. All variables were assessed with Shapiro-Wilk test and visual histogram check for normality. All zip codes with at least 1 case or 1 death were included in the respective models. Non-normal variables including the dependent variables above were log transformed. Interactions were assessed in the first adjusted model and were dropped after a likelihood ratio test. Multicollinearity was addressed by performing ridge regression technique where the ridge parameter was adjusted at 0.004 after root mean square error comparison. The final linear regression model presented the log transformed dependent and independent variables, the interpretation was presented as percent change in independent variable required for 1% change in number of cases or deaths. All inferential linear model designs were presented in **supplemental table 3**.

Sensitivity analysis for all positive cases was performed by adding comorbidities of hypertension and cancer was performed. Additionally, similar analysis for comorbidities of diabetes and asthma as described above was performed for zip codes in Chicago and New York. Additional sensitivity analysis was done for zip codes above median number of cases and deaths for the city/county of origin of the zip code.

Alpha for the study was set at 0.01 and analysis was performed in SAS 9.4 Cary NC.

## Results

Overall, as of 04/11/2020 there were 442 zip codes reporting 93,170 positive COVID-19 cases. The range of cases spanned from 0 to 1,728 per zip code. Only the zip codes in Oakland county of Detroit and King’s County in Seattle reported mortality. Overall, 138 zip codes reported mortality ranging from 0 to 25 dead.

Among the 442 zip codes studied the median age was 38.2 (34.8 - 42) years, 51% (50% - 53%) were female, 45% of zip code residents were white (13% - 70%), 6% of zip code residents were African American (2% - 21%) asnd 15% of zip code residents were Hispanic (6% - 40%). Other demographics of the 443 zip codes are presented in **table 1**.

**Table 1:**
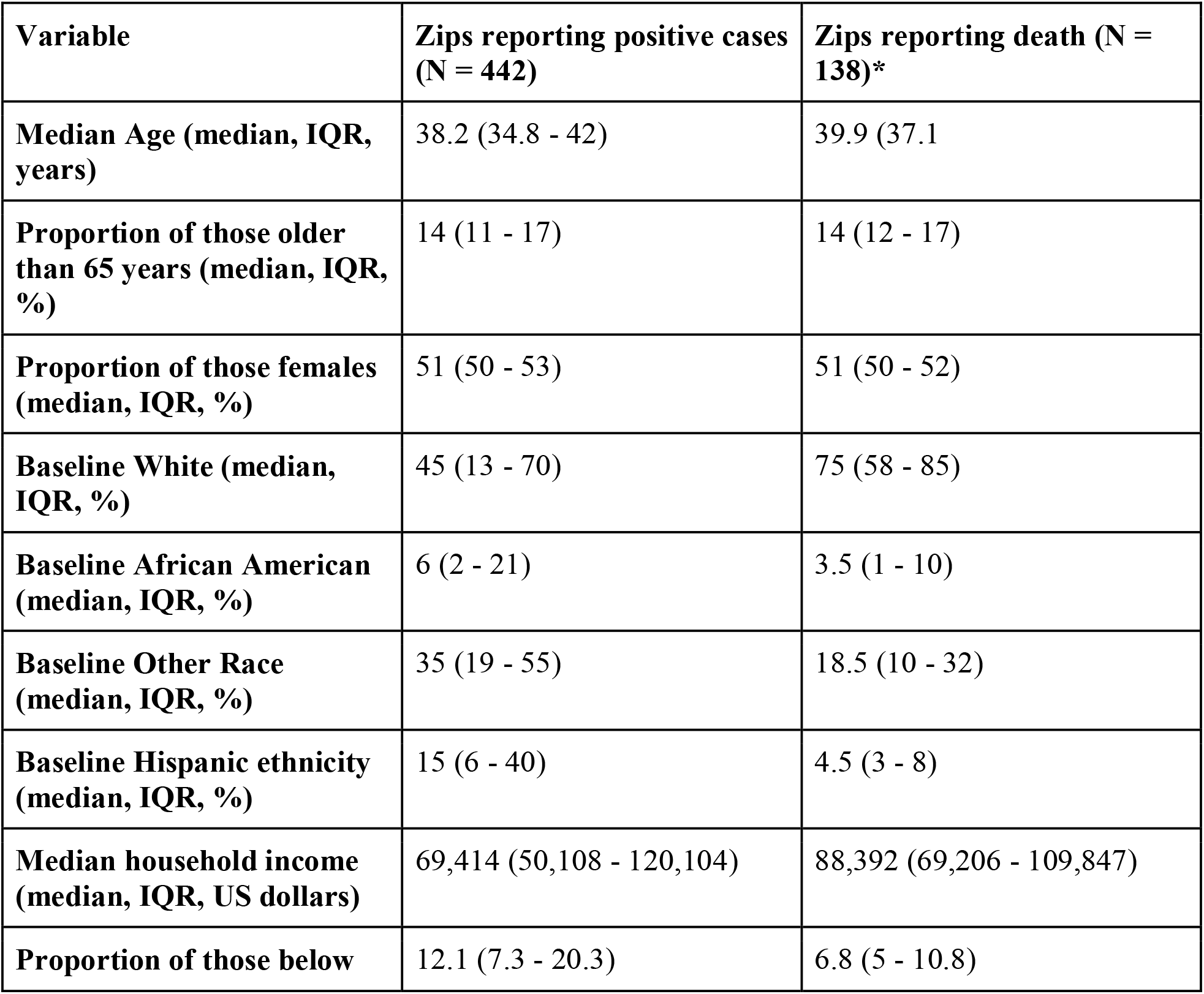
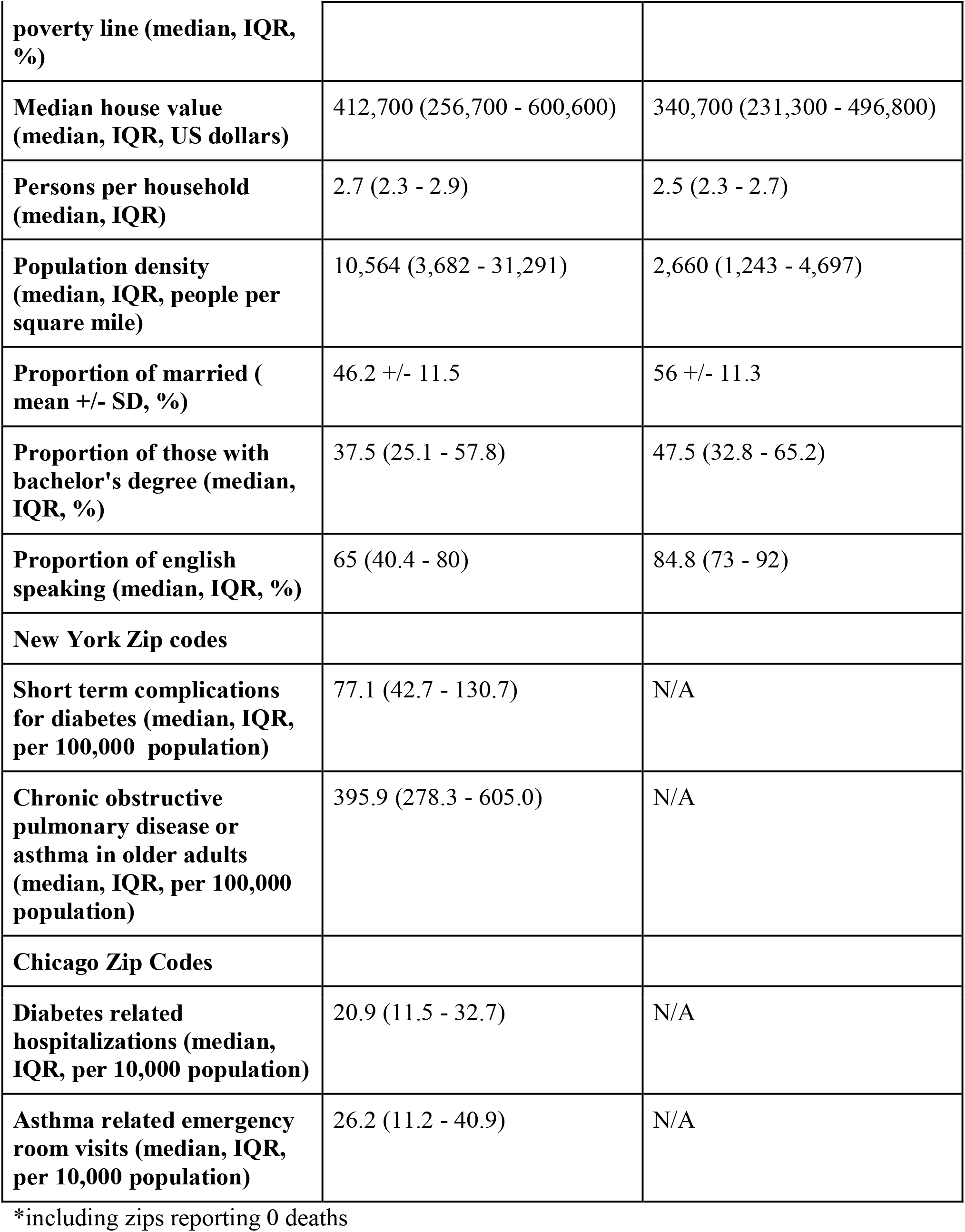
Baseline demographics of zip codes used in the study (current as of 4/11/2020)

### Modelling for positive cases

In a multivariable linear regression model (**table 2**) it was noted than proportion of residents above the age of 65 years, proportion of African American residents, proportion of females, persons per household and population density of the zip code was significantly associated with increased likelihood of positive cases in a zip code (P <= 0.01).

In a sensitivity analysis using dichotomous hypertension and cancer county level data the associations noted above remain (**table 2**). On further sensitivity analysis of NYC and Chicago zip codes it was clear that proportion of residents above the age of 65 and population density continued to be significantly associated with increased likelihood of positive cases. Among the zip codes with greater than median number of cases (129), only population density was associated with increase in case. For 1% increase in population density there was 0.27% (0.13% - 0.40%) increase in number of cases (P = 0.0002). All the 4 models mentioned here are presented in supplemental table 3.

**Table 2.**
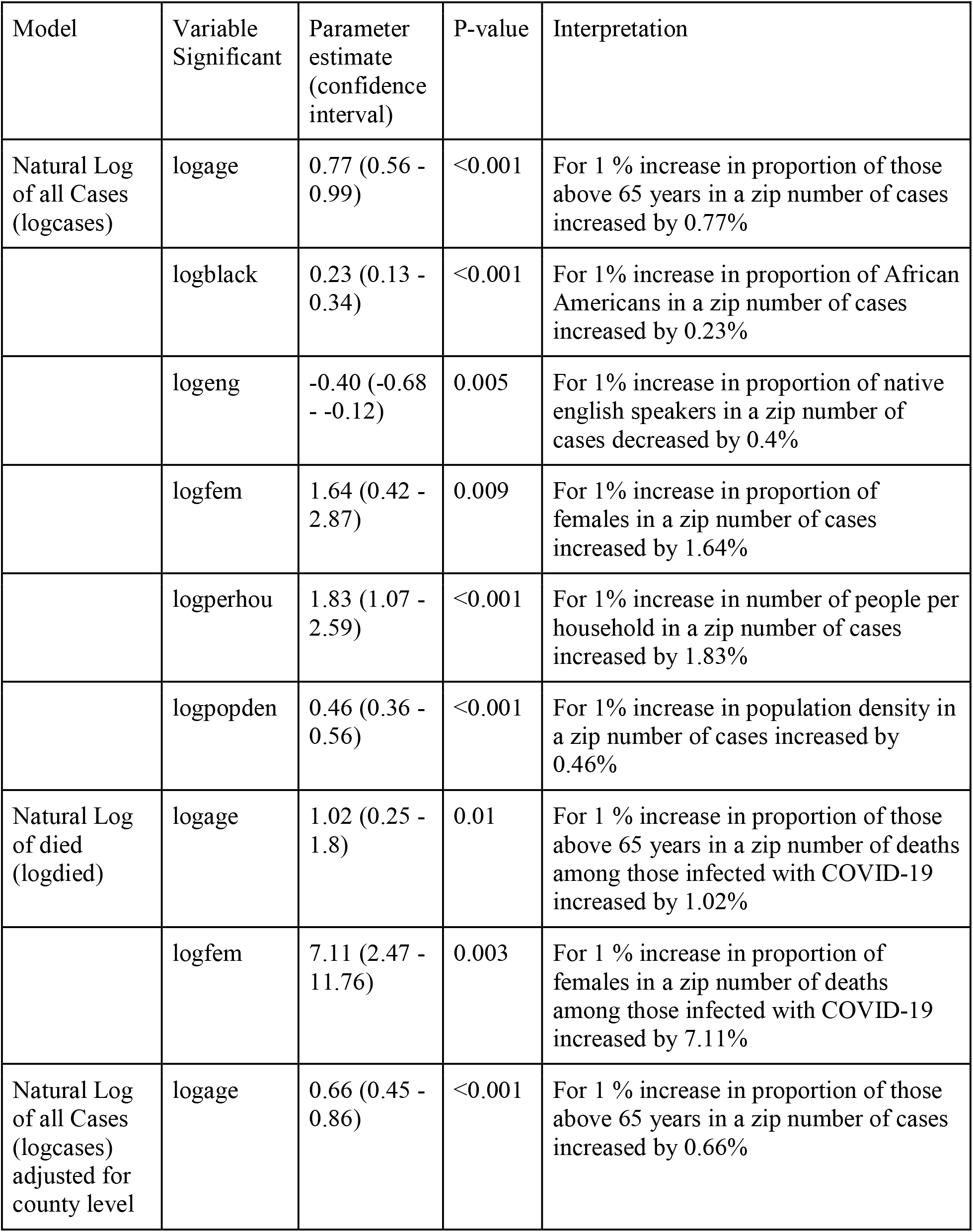
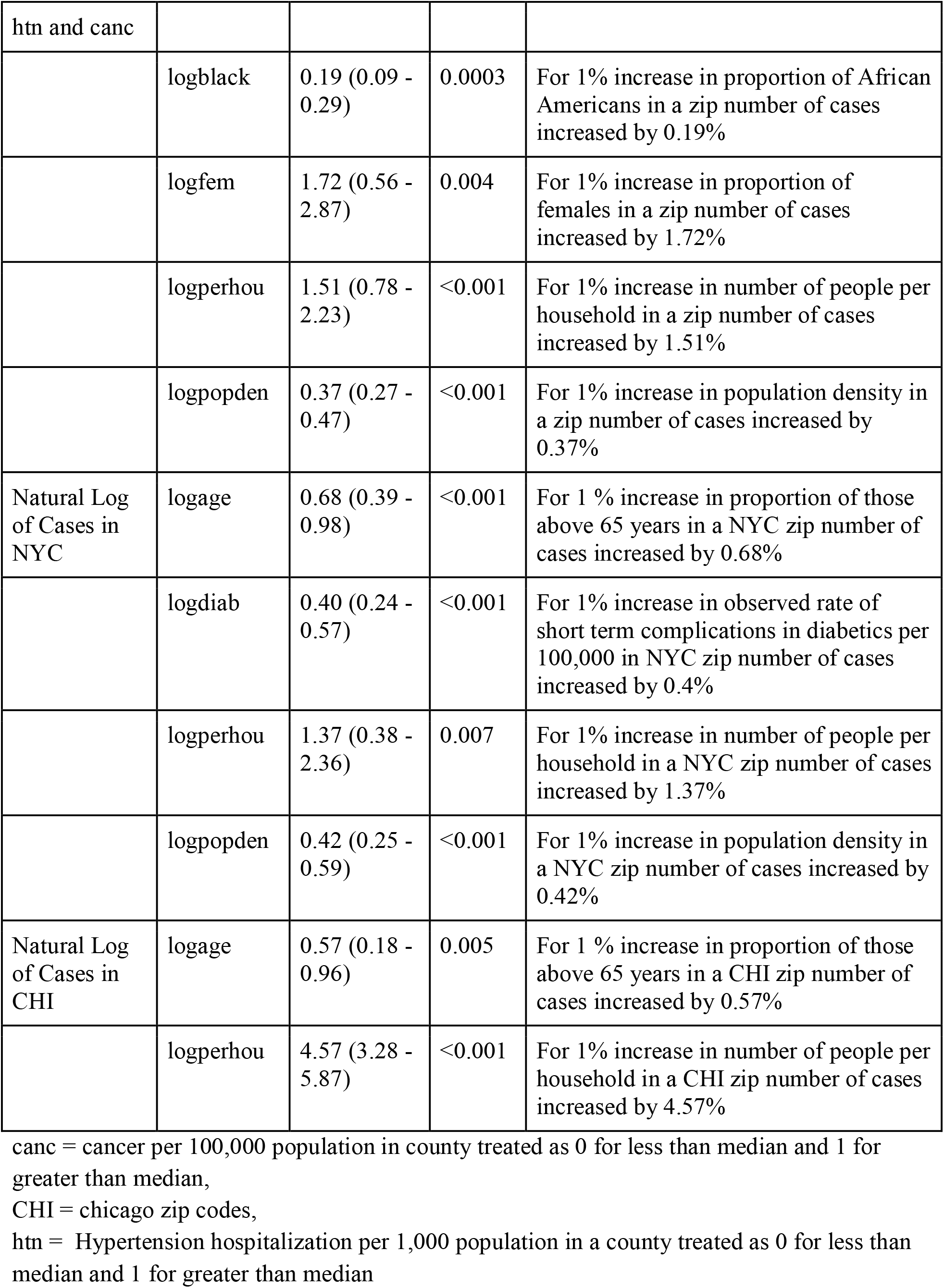
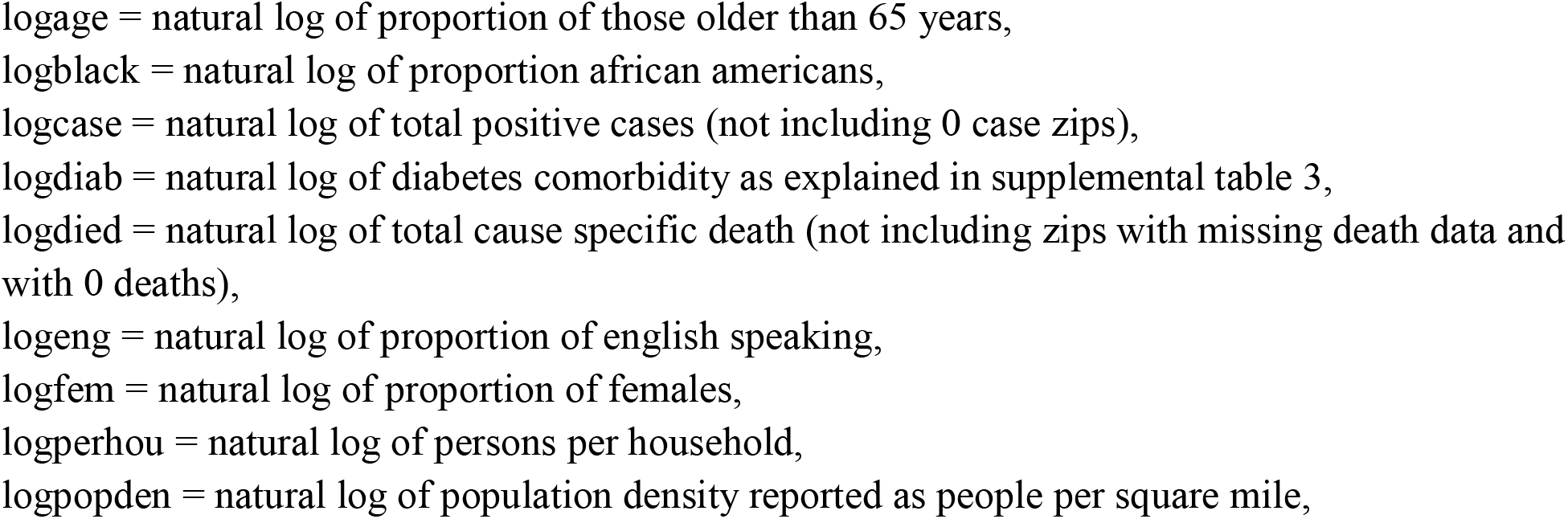
Linear Regression Modeling Specific Results of significant variables. All proportions presented are multivariable with models presented in supplemental table 3

### Modelling for mortality

Moreover, in a multivariable linear regression model (**table 2**) proportion of residents above the age of 65 years, and proportion of females in a zip code was associated with increased likelihood of COVID-19 related mortality in a zip code (P <= 0.01). However, there was no such association noted in zip codes with greater than median number of deaths (2).

## Discussion

Our findings hold a mirror revealing yet another example of social inequities and disease. This study evaluated COVID-19 cases and mortality rates, integrating the data with reported socioeconomic measures by zip code. We demonstrate that zip codes with a greater proportion of residents above 65 years of age, African American residents, persons per household, and population density were significantly associated with an increased likelihood of COVID-19 cases. After sensitivity analysis, population density remained the factor most significantly associated with COVID-19 cases. These results highlight the importance of social distancing or lack thereof in poorer neighborhoods while giving early signals of racial inequalities which has been noted in other infectious disease and chronic disease epidemiology.

The COVID-19 related health disparities and inequities are not entirely unfamiliar to health practitioners in the U.S. The HIV epidemic remains one of the greatest public health challenges faced in the modern world. In survival analyses, a racial divide was demonstrated in HIV outcomes, with an increasingly disproportionate burden borne by African Americans (7). Additionally, reports of socioeconomic disparities and HIV prevalence has been well reported, with associations including age, wealth, household expenditure, and rural versus urban residence (4, 8). Like other preliminary reports, our analysis of the COVID-19 pandemic has also revealed age and race to be positive predictors in COVID-19 cases (6, 9). However, when biologic factors such as age and race were held constant, like the HIV epidemic, we found other significant variables in the structural environment that impacted the rate of COVID-19 cases. The impact of socioeconomic disparities on health has been well documented and established, revealing that those with the lowest income and who were least educated were consistently least healthy (10, 11), with correlations present down to the zip code level (12). Our results suggest that even though race, age, and gender were noted initially, zip code level population density is most significant.

There’s a correlation between overcrowding and the historic spread of infectious disease. The influenza pandemic of 1918, popularly known as the Spanish Flu, killed more than 50 million people worldwide (13). Analysis of the specific factors that made that influenza especially catastrophic was overcrowding, which increased both risk of flu by 10-fold and severity by 5-fold (14). In fact, records from an “open air” hospital in Boston, Massachusetts found that a combination of fresh air, sunlight, and reusable face masks substantially reduced deaths among some patients and new infections among medical staff (15). Social distancing has remained an important way of combating the spread of COVID-19. To date, in the United States, all 50 governors have declared states of health emergencies. Officials have implemented recommendations around social distancing, which includes the closing of nonessential businesses, closure of schools, and the implementation of stay at home orders (16). Targeted social distancing has been established as an effective way to reduce viral transmission, with a previous study of the 1957-58 influenzas showing a reduction in attack rate by >90% (17). However, social distancing is more difficult for some populations than others. Our data demonstrate that among the zip codes with the highest population density, COVID-19 cases were greater. This is unsurprising and likely due to the historical impact of population density and overcrowding observed in past pandemics.

There are certain limitations which should be considered while considering the aforementioned results. The study results have potentially changed in terms of reported number of cases and deaths however, the models generated, and effects described are expected to potentially remain the same given the overall power of the study and multiple sensitivity analysis performed. Even though multiple socio-demographic variables were utilized, the authors acknowledge that this study should not be interpreted out of context of currently available data. Specifically, chronic conditions increase the risk of acquiring and dying from COVID-19 and in this study such adjustment was limited from such adjustments. Another metric about the number of positive cases may point towards discrepant testing/swab availability. However, the authors feel that socio-demographic factors may be responsible for such discrepancy in testing and would contribute to multicollinearity and not change the effect noted above.

## Conclusion

Race has long been used as a proxy for socioeconomic status, with growing pressure to specifically name the variables truly at play. In this analysis, even though signals regarding the role of race and gender were noted initially, after sensitivity analysis, we found that the most significant predictor of a positive COVID-19 case was the structural environment of an individual, namely, the population density within their zip code. This reality makes preventative measures, such as social distancing policies, difficult to implement. This research highlights the need for community-specific outreach and interventions to address the COVID-19 pandemic.

## Data Availability

This study was performed using publicly available data and complies with the Declaration of Helsinki, however given the public nature of the data locally appointed ethics committee approval and informed consent was not required.

